# Effects of successful percutaneous coronary intervention of chronic total occlusions with demonstrable viability or ischemia: A protocol for a systematic review

**DOI:** 10.1101/2023.08.01.23293494

**Authors:** Luís Leite, Tomás Carlos, Gustavo Campos, Helena Donato, Rodolfo Silva, Antero Abrunhosa, Lino Gonçalves, Maria João Ferreira

## Abstract

**Background:** Chronic coronary total occlusion (CTO) is a common finding in patients referred to coronary angiography. To sustain blood flow distally to the occlusion site, collateral circulation is formed from preexisting vessels. While these collaterals may partial or completely preserve perfusion at rest, they may be insufficient when increased blood flow is needed. According to current guidelines, percutaneous coronary intervention (PCI) is recommended solely in patients with resistant angina despite optimal medical therapy or when a large area of documented ischemia in the territory of the occluded vessel is present. Randomised controlled trials (RCTs) suggest its benefit in the improvement of patients’ quality of life and symptoms, despite their conflicting results concerning prognosis and left ventricular function. However, most of these studies often lack data regarding myocardial ischemic burden and viability, assessed by imaging methods. Therefore, the purpose of the systematic review is to comprise and analyse the literature on whether viability or ischemia-guided PCI of CTO, identified by imaging methods, has an impact on the clinical outcomes of the patients.

**Methods:** We will conduct a thorough research in different databases, including PubMed/MEDLINE, EMBASE, Cochrane Central Register of Controlled Trials (CENTRAL), Web of Science Core Collection, Clinical trials in the European Union, and ClinicalTrials.gov. We will include RCTs, cohort studies, cross-sectional studies, and case-control studies, in which patients with established CTO were tested for viability and/or ischemia before the decision to perform PCI and were evaluated by post-intervention testing or clinical endpoint follow-up. Two authors will independently review the selected studies, and any discrepancies will be solved by a third element. Subsequently, data from the various eligible studies will be extracted and analysed by two different authors. No subgroup analysis is planned.

**Ethics and disclosure:** Ethical approval will not be required for this study as it is an analysis of previously published articles (i.e., secondary data). The results will be published in a peer-reviewed journal.

**PROSPERO registration:** CRD42023426858.

## INTRODUCTION

Chronic total coronary occlusions (CTO) are defined as angiographic evidence of complete vessel occlusion with Thrombolysis in Myocardial Infarction (TIMI) flow grade 0 within an epicardial coronary artery, and an estimated duration of at least three months [1]. Its prevalence is not entirely established, with reporting rates varying from 16%-18% [2] to approximately 30% [3,4] in patients referred for coronary angiography. Notably, within the subgroup diagnosed with stable angina pectoris, it is believed that up to one fourth of this population exhibit this type of coronary disease [5].

In the presence of a CTO, blood supply to the myocardium is solely from collateral vessels. The development of a collateral circulation network is a complex physiologic response induced by the chronic state of hypoxia, by the pressure gradient created, as well as by several different humoral factors. There is an individual variation on the ability to develop well-formed collaterals that may avoid a transmural myocardial infarction and preserve ventricular function. However, it is widely believed that, although the collateral network may have the ability to preserve perfusion at rest, it could be insufficient when increased blood flow is needed [4].

Moreover, the chronic supply defect from the main artery may cause myocardium hibernation, characterized by reversible reduced regional contractile function that could partial or completely recover after reperfusion or revascularization [6,7]. Bearing this in mind, it would be reasonable to refer these patients to percutaneous coronary intervention (PCI). Nonetheless, current guidelines, prevailing since 2018, recommend CTO PCI only in patients with angina resistant to optimal medical therapy or with a large area of documented ischemia in the territory of the occluded vessel (with a class of evidence IIa) [8].

Until recently, there was reluctance in attempting PCI in CTO patients, with solely an estimate 10%-15% of all eligible CTO lesions being referred [9,10]. This was based on lower procedural success and high rate of complications, such as urgent coronary artery bypass graft (CABG) surgery, coronary perforation, acute reocclusion, collateral loss and major adverse cardiac events, including death [11]. During the past decade, important developments in interventional materials and techniques have led to the achievement of high rates of procedural success, defined as a residual stenosis <30% with antegrade TIMI flow grade 3 in the CTO target vessel [12,13]. Indeed, recent studies such as the EXPLORE trial, suggested that PCI of CTO was safe even in patients with a concurrent ST-elevation myocardial infarction (STEMI) in other main artery, with no periprocedural death or emergency CABG [14]. Also, evidence suggests that the higher mortality and morbidity found in patients with multivessel disease, in comparison to single-vessel disease, might be partially attributed to the presence of concomitant CTO, contributing to the declining of LV function [15].

Hence, it urges to clearly define which patients are candidates for PCI on top of optimal medical therapy (OMT), since currently we have no accurate method that predicts who will benefit from that [16]. In the past few years, randomized clinical trials (RCTs) have focused on evaluating whether CTO-PCI in addition to OMT improved patients’ symptoms and prognosis. Overall, it is generally accepted that it improves patients’ quality of life and symptoms, mainly angina and physical exercise limitation, when compared to no PCI [16–20]. DECISION-CTO trial was one of the few studies that did not show this relative benefit [21]. However, in this study, revascularization of non-CTO lesions was performed after randomization in OMT+CTO-PCI or solely OMT, potentially contributing to the absence of differences [20]. When prognosis and LV function are concerned, studies show more conflicting results. In a subgroup analysis of the EXPLORE trial, Elias *et al*. demonstrated that in the dysfunctional segments afflicted by the chronic occlusion, PCI was associated with marked improvement of regional function, measured by cardiac magnetic resonance imaging (cMRI), especially if transmural extent of infarction (TEI) was lower than 50% [22]. Different research using cMRI suggested that in segments with TEI below 75% there was a tendency for improvement in segmental wall thickening (SWT), and that the extent of improvement in LV ejection fraction (LVEF) correlated with the size of infarcted area before procedure [23]. However, mean LVEF remained practically unchanged, with the same finding being observed in the COMET-CTO trial [20]. Furthermore, REVASC trial showed no differences in regional wall motion of the myocardium affected by the CTO lesions after PCI [24]. Moreover, IMPACTOR-CTO trial evaluated the impact of PCI on the composite endpoint of all-cause death, myocardial infarction, and unplanned revascularization six months after the procedure, with no significant differences described [17]. Lastly, OAT-trial demonstrated a tendency for reinfarction in the PCI group, at a follow-up time of approximately four years [25].

In the published RCTs there are very few data regarding myocardial ischemic burden and viability, often assessed by imaging methods, such as cMRI, single-photon emission computed tomography (SPECT), stress echocardiography or positron emission tomography (PET). Currently, ISCHEMIA-CTO trial is underway to evaluate whether CTO-PCI improves quality of life and major adverse cardiovascular events in patients with significant ischemic burden [26].

To the best of our knowledge, a systematic review of the literature on whether viability or ischemia-guided PCI of CTO have an impact in clinical outcomes is yet to be conducted. We aim to address this subject, evaluating all-cause mortality, nonfatal myocardial infarction, symptomatic improvement, and ischemic burden reduction in the follow-up of patients with documented viability or ischemia previously to the decision to perform PCI.

## METHODS AND ANALYSIS

The protocol and completed systematic review will be reported following the Preferred Reporting Items for Systematic Reviews and Meta-analyses (PRISMA) Protocols and the Prisma checklists, respectively.

### Eligibility criteria

In this systematic review, our objective is to identify RCTs, cohort studies (prospective or retrospective in nature), cross-sectional studies, and case-control studies, in which patients with established CTO were tested for viability and/or ischemia before the decision to perform PCI and were evaluated by post-intervention testing or clinical endpoint follow-up. The diverse testing modalities encompassed cMRI, PET, SPECT or stress echocardiography. Studies lacking viability and/or ischemia testing before PCI or lacking measured clinical endpoints or post-intervention testing will be excluded. Table 1 presents our PICO description. If the same study is represented in different articles, the one with the largest sample size or more suitable data for our specific aim will be selected. No restriction regarding publication year will be set; therefore, we will be including studies from inception to the 31st of October 2023. There will be no restrictions on publication status or language of publication. We will resort to a professional translator if needed.

**Table 1.** PICO description.

| Abbreviation | PICO | Elements |
| --- | --- | --- |
| P | Population | Patients with a CTO who had performed viability or ischemia exams before the decision to perform PCI, with a post-intervention clinical evaluation and/or cardiac testing (LV function, ischemia burden evaluation) |
| I | Intervention | Successful PCI with documented viability or ischemia |
| C | Comparator | Patients with a CTO who had no demonstrable viability or ischemia before performing successful PCI, had |
|  |  | demonstrable viability or ischemia but have not performed<br>PCI or in whom PCI was unsuccessful |
| O | Outcome | All-cause mortality, nonfatal myocardial infarction,<br>symptomatic improvement, left ventricular function<br>improvement, and ischemic burden reduction |

### Information sources

We will comprise literature published in the following databases: PubMed/MEDLINE, EMBASE, Cochrane Central Register of Controlled Trials (CENTRAL), Web of Science Core Collection, and we will also consult Clinical trials in the European Union, and ClinicalTrials.gov to check for ongoing or unpublished trials. If any relevant unpublished trial is found, the corresponding author listed will be contacted to grant access to the required information. If no response is given or the author decides not to share the data, this will be listed as the reason for excluding such a trial Grey literature will also be queried to include all possible articles on the subject. We will also hand search in the reference list of the included articles. No pre-prints will be included.

### Search strategy

Detailed search strategies will be conducted in the previously described databases, in consultation with a medical librarian with expertise in systematic review searching. The search strategy will contain controlled vocabulary (e.g., Medical Subject Headings (MeSH)) and text words searches adapted to each one of the databases regarding its own special requirements. To merge the search terms effectively, Boolean operators “AND” and “OR” will be used. An example of the search strategy used in PubMed/MEDLINE is depicted in Table 2.

**Table 2.** Search strategy.

| PubMed/MEDLINE |  |
| --- | --- |
| Query | Search |
| #1 | (Coronary Occlusion[Title/Abstract]) AND (chronic[Title/Abstract])) OR (((<br>"Coronary Occlusion/diagnosis"[Mesh] OR "Coronary Occlusion/diagnostic<br>imaging"[Mesh] OR "Coronary Occlusion/therapy"[Mesh] )) AND "Chronic<br>Disease"[Mesh]) |
| #2 | ("Heart/diagnostic imaging"[Mesh] OR "Cardiac Imaging Techniques"[Mesh] OR<br>"Ventricular Function, Left"[Mesh] OR "Positron-Emission Tomography"[Mesh]<br>OR "Magnetic Resonance Imaging, Cine"[Mesh] OR "Adenosine"[Mesh] OR<br>"Contrast Media"[Mesh] OR "Dobutamine"[Mesh] OR "Gadolinium"[Mesh] OR<br>"Echocardiography"[Mesh] OR "Fibrosis"[Mesh] OR "Necrosis"[Mesh] OR<br>"Myocardial Ischemia"[Mesh] OR "Myocardial Stunning"[Mesh] OR "Ventricular<br>Dysfunction, Left"[Mesh]) OR (Cardiac Imaging Techniques[Title/Abstract] OR<br>Techniques, Intracardiac Imaging[Title/Abstract] OR Ventricular Function,<br>Left[Title/Abstract] OR Myocardial Perfusion Imaging[Title/Abstract] OR<br>Positron-Emission Tomography[Title/Abstract] OR Positron Emission<br>Tomography Computed Tomography[Title/Abstract] OR Magnetic Resonance<br>Imaging Cine[Title/Abstract] OR Adenosine[Title/Abstract] OR Contrast<br>Media[Title/Abstract] OR Dobutamine[Title/Abstract] OR<br>Gadolinium[Title/Abstract] OR Echocardiography[Title/Abstract] OR<br>Echocardiography, Stress[Title/Abstract] OR Fibrosis[Title/Abstract] OR<br>Necrosis[Title/Abstract] OR Myocardial Ischemia[Title/Abstract] OR Ventricular<br>Dysfunction, Left[Title/Abstract] OR Myocardial Stunning[Title/Abstract] OR<br>Myocardial hibernation[Title/Abstract]) |
| #3 | (Percutaneous Coronary Intervention[Title/Abstract] OR Angioplasty, Balloon, Coronary[Title/Abstract] OR Atherectomy, Coronary[Title/Abstract] OR Coronary Revascularization, Percutaneous[Title/Abstract] OR Myocardial Revascularization[Title/Abstract] OR "percutaneous coronary intervention"[MeSH Terms] OR "Myocardial Revascularization"[Mesh] ) |
| #4 | Search #1 AND #2 AND #3 |

### Data management

All studies and literature reviewed will be imported to EndNote citation software and all duplicates will be removed.

### Selection process

Two independent reviewers will conduct the selection phase of the studies during the month of January 2024. Any discrepancy found between the two reviewers will be solved by a third reviewer. The remaining studies will be full-text reviewed and included or excluded according to our predefined eligibility criteria. A flow chart similar to Fig. 1 will be presented to detail the study selection process for this review.

**Fig. 1.**
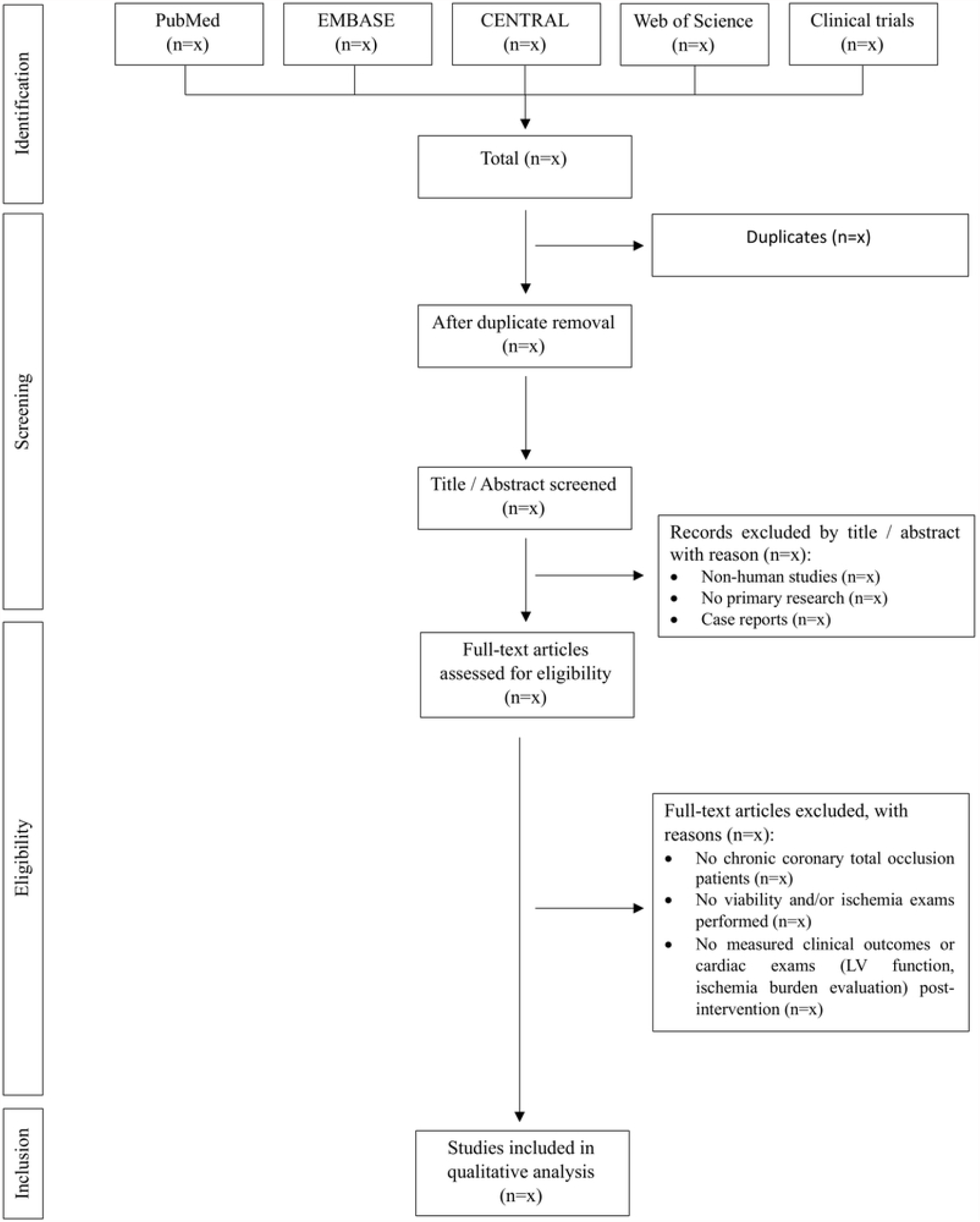
Flowchart diagram presenting the selection process for the studies.

### Data collection process

Data collection will be performed after the selection process. Two independent reviewers will extract the relevant data for our systematic review from the various eligible studies. A third reviewer will solve any found discrepancies and validate the other results.

### Data items

Collected data from the obtained studies will encompass authors’ names, sample size, study design, number of eligible patients, average age, gender distribution, type of cardiac exam conducted pre and post intervention, decision to perform PCI or solely OMT, success of PCI, definition of myocardial ischemia and viability assessment for each study and type of clinical follow-up performed. Reasons for the exclusion of full text records will be reported.

Details on the selection process of the studies and included patients will be documented into a flow chart, following PRISMA checklists.

### Outcomes

Our systematic review aims to assess whether viability or ischemia-guided PCI of CTO have an impact on clinical outcomes. To achieve this, we will analyse diverse parameters across the different studies such as all-cause mortality, nonfatal myocardial infarction, symptomatic improvement, left ventricular improvement and ischemic burden reduction post PCI (successful vs non successful) or solely OMT. No subgroup analysis is planned at first hand; however, it may be performed if motivated by the literature review.

### Risk of bias

Risk of bias of individual studies will be assessed independently by two reviewers, with a third reviewer assisting with any disagreements. Since we will be including diverse types of studies, we will use different tools to identify the risk of bias depending on each study’s characteristics.

We will use Cochrane Risk of Bias 2.0 –tool for assessing risk of bias in randomized trials, and ROBINS-I tool - Risk Of Bias In Non-randomized Studies - of Interventions.

### Data synthesis

We will provide a narrative synthesis of the findings from the selected studies, structured around the viability and/or ischemia exam, and type of clinical outcome measured.

### Confidence in cumulative evidence

The strength of the body evidence will be assessed using the Grading Recommendations Assessment, Development and Evaluation (GRADE).

### Patient and public involvement

This is a protocol for a systematic review based on previously published data. No participant recruitment or involvement will be necessary.

### Ethics and dissemination

This systematic review will be conducted upon public and already published data; therefore, no ethical approval is necessary. The resulting work will be published in a peer-reviewed journal and presented at relevant conferences. Any necessary modifications of the protocol will be dated and reported through a detailed description of the rationale for the adjustments.

## DISCUSSION

Even though CTO are a common finding in patients with coronary artery disease, there are conflicting results regarding the benefit of performing PCI in these lesions. Most clinical studies lack data regarding myocardial ischemia burden and viability documentation, therefore a systematic review on the clinical impact of ischemia/viability-guided CTO PCI is needed.

This study has limitations as it is a secondary study dependent on the published data. Moreover, there are several imaging modalities to diagnose ischemia or viability, each one with its own criteria. On the other hand, the clinical impact of the intervention is measured with different outcomes within each study, which could make the discussion more demanding to perform. Therefore, we will provide a narrative synthesis of the findings, structured around the diagnostic exams and type of clinical outcome.

## SUPPORTING INFORMATION

S1 Checklist. PRISMA-P (Preferred Reporting Items for Systematic review and Meta-Analysis Protocols) 2015 checklist: Recommended items to address in a systematic review protocol*.

## AUTHOR CONTRIBUTIONS

Conceptualization: Luís Leite, Tomás Carlos, Maria João Ferreira.

Investigation: Luís Leite, Tomás Carlos, Gustavo Campos, Lino Gonçalves, Maria João Ferreira.

Methodology: Luís Leite, Tomás Carlos, Gustavo Campos, Helena Donato.

Supervision: Maria João Ferreira.

Writing – original draft: Luís Leite, Tomás Carlos, Gustavo Campos.

Writing – review & editing: Luís Leite, Tomás Carlos, Gustavo Campos, Helena Donato, Rodolfo Silva, Antero Abrunhosa, Lino Gonçalves, Maria João Ferreira.

## Data Availability Statement

No datasets were generated or analyzed during the current study. All relevant data from this study will be made available upon study completion.

## Funding

The authors received no specific funding for this work.

## Competing interests

The authors have declared that no competing interests exist.

## Notes

### Competing Interest Statement

The authors have declared no competing interest.

### Funding Statement

The author(s) received no specific funding for this work.

### Author Declarations

This systematic review will be conducted upon public and already published data therefore, no ethical approval is necessary.

